# LiMeTrack: A lightweight biosample management platform for the multicenter SATURN3 consortium

**DOI:** 10.1101/2025.09.01.25334868

**Authors:** Florian Heyl, Jonas Gassenschmidt, Lukas Heine, Frederik Voigt, Jens Kleesiek, Oliver Stegle, Jens T. Siveke, Melanie Boerries, Roland F. Schwarz, Laura Godfrey

**Author notes:** equally contributing first author (Florian Heyl, Jonas Gassenschmidt, Lukas Heine, Frederik Voigt).

## Abstract

Biomedical research projects involving large patient cohorts are increasingly complex, both in terms of data modalities and number of samples. Such projects require robust data management solutions to foster data integrity, reproducibility and secondary use compliant with the FAIR (Findability, Accessibility, Interoperability, and Reusability) principles. SATURN3, a national German consortium with 17 project partner sites investigates intratumoral heterogeneity in three hard-to-treat cancers using patient biosamples actively collected at six project centers. The complex multicenter workflow of SATURN3 thereby includes multiple analyte types across bulk and single-cell sequencing, and spatial omics technologies.

To avoid the risk of data loss and de-synchronization across the partner sites, metadata harmonization and documentation in a central infrastructure is essential. In addition, real-time monitoring of the processing status of the samples must be transparently accessible to all project partners throughout the project. This use case goes far beyond the documentation capabilities of spreadsheets or locally shared files that are susceptible to security vulnerabilities, versioning mistakes, data loss and type conversion errors. Existing data management tools are often too complex to set up or lack the necessary flexibility to be adopted for specific project needs.

To address these challenges, we introduce LiMeTrack, an online biosample management platform with customizable and user-friendly data entry schemes and a real-time dashboard for providing an overview of project and sample status. LiMeTrack simplifies the creation and export of standardized sample sheets, streamlining subsequent bioinformatics analyses and biomedical research workflows. By integrating real-time monitoring with robust sample tracking and data management, LiMeTrack improves research transparency and reproducibility, ensures data integrity and optimizes workflows, making it a powerful solution for sample management in modern multicenter biomedical research scenarios.

## Introduction

Research projects involving patient biosamples are becoming increasingly complex. Larger patient numbers are often achieved through multicenter recruitment, while data layers become increasingly multi-modal, combining multi-omics, imaging technologies and patient-derived cell systems. In this environment, a dedicated data management strategy and platform lay the foundation for a successful and reliable research project with a trustworthy data basis and reproducible analysis workflows.

The SATURN3 project is a national German multicenter cancer genomics consortium investigating temporal and spatial heterogeneity in three hard-to-treat cancers across 17 partner sites (saturn3.org). Funded by the Federal Ministry of Research, Technology and Space (BMFTR) SATURN3 is part of the National Decade against Cancer initiative and one of the largest such consortia in Germany. Out of the 17 partner sites, six actively recruit patients and collect biomaterial (tissue, blood) that is processed in a complex, multimodal workflow. Overall, SATURN3 aims to recruit 150 patients donating more than 800 biosamples with temporal and spatial resolution. Processing involves pathological assessment of tumor cell content and tissue quality, followed by diverse multi-omics analyses including bulk and single-cell sequencing, and spatial transcriptomics. Wet lab and bioinformatics analyses are often carried out in different laboratories or core facilities at separate partner sites.

Consequently, SATURN3 poses distinct challenges to research data management and biosample tracking. To prevent miscommunication, data loss and desynchronization a central infrastructure platform must provide simultaneous real-time access to all samples processed at all partner sites, while catering to the requirements of diverse data modalities and the different stakeholders with varying levels of technical expertise. These demands go far beyond the documentation capabilities of spreadsheets or shared files that are susceptible to security vulnerabilities, incomplete datasets, data loss and type conversion errors [1]. For example, clinical project partners involved in patient recruitment rely on an intuitive graphical user interface (GUI) for entering clinical metadata that is accessible and usable without prior training or experience. Errors and inconsistencies must be recognized and flagged at the earliest stage of data entry, for both individual and batch input. Wet lab personnel and laboratory core facilities handling biosamples must be able to dynamically document sample intake and processing status efficiently across multiple samples. Metadata and status information must be available to all partners in real time during the course of the project. Data analysts and bioinformaticians on the other hand require the export of custom sample sheets for downstream data analyses specific to the bioinformatics workflows conducted. These sample sheets must be integrity checked and be available in a structured and machine readable format adapted to the specific requirements and needed covariates for the analyses conducted. Ultimately, at the project management stage data security and consistency are paramount and require a dedicated user management system. Read and write permissions should be set on an individual and group level basis using an intuitive user interface without needing programming skills. In addition, all data must be exportable in a structured format to allow easy integration into platforms such as the German Human Genome Phenome Archive (GHGA [2,3], and European Genome Archive (EGA), which facilitate a controlled secondary use of human patient data in line with the FAIR principles (Findable, Accessible, Interoperable and Reusable) [4].

While several platforms for biosample management exist, they often focus on standardised biobanking workflows [5], are closed source [6,7] or are too complex to adapt to specific needs of an individual research project in a limited amount of time [8,9]. For example, REDCap (Research Electronic Data Capture) is widely used in clinical and epidemiological studies and offers robust form-based data entry with built-in validation and audit trails [9]. However, it implements a rigid data model and has limited support for real-time sample tracking, interactive visualization, or downstream analysis. Moreover, using REDCap requires an organizational membership in the REDCap consortium which limits modification and deployment outside of institutional agreements. Similarly, SODAR (System for Omics Data Access and Retrieval) provides a powerful framework for managing multi-omics metadata and structured experiment annotation based on ISA-Tab [8]. While open-source and well-suited for institutional-scale omics data organization, SODAR is aimed at joint storage of primary and meta data, optimized to work within the Berlin Institute of Health infrastructure and thus requires significant technical expertise for setup and customization. FAIRDOM-SEEK is a community-built platform covering a broad range of metadata annotation and semantic integration within the life sciences domain [10]. However, its primary focus is on the organization and dissemination of structured datasets and computational models rather than on real-time sample tracking, biosample logistics, or management of dynamic sample workflows.

To fill the aforementioned gaps for the national SATURN3 consortium we have developed the lightweight metadata and biosample tracking platform LiMeTrack.

LiMeTrack serves as an all-in-one solution for biomedical research projects involving patient biosamples, wet lab experiments and sequencing analyses (Figure 1). It is especially designed to be lightweight and flexible, requiring minimal programming skills for data entry, and provides real-time monitoring and sample tracking throughout the project. Customizable and flexible data exports ease downstream analyses of all derived data. Using LiMeTrack, small and big teams alike can be equipped with a flexible and robust data management platform for tracking metadata, sample processing status and data availability.

**Figure 1:**
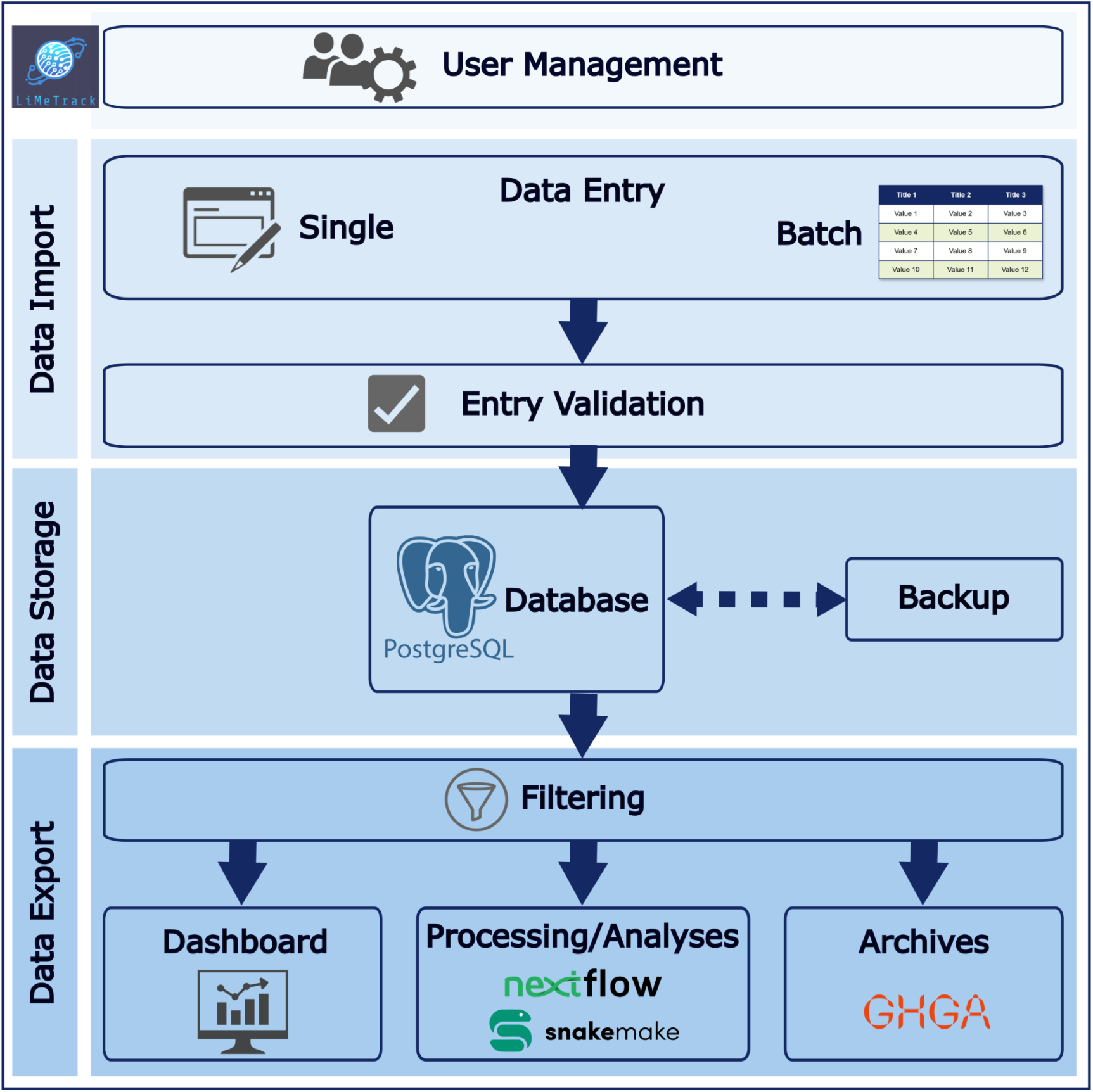
Overview of the architecture and features of LiMeTrack.

## Results

The LiMeTrack frontend is built on four key interactive elements that can also be utilized by untrained personnel through an intuitive graphical user interface (GUI) (Figure 2a): a *Privilege Module* in which administrators can define user and group-specific privileges and access rights; a *Project Module*, which summarises key information about the administered project, related tools and websites, and also acts as the UI’s landing page; a *Sample Module* with a dedicated *Sample Form* for entering project-specific sample metadata and parameters manually or through batch import and a *Sample Overview* listing with customisable filters which shows information for all samples; and finally a real-time *Visualization Module* termed *Dashboard*, which provides an overview of all samples at each processing stage of the project.

**Figure 2:**
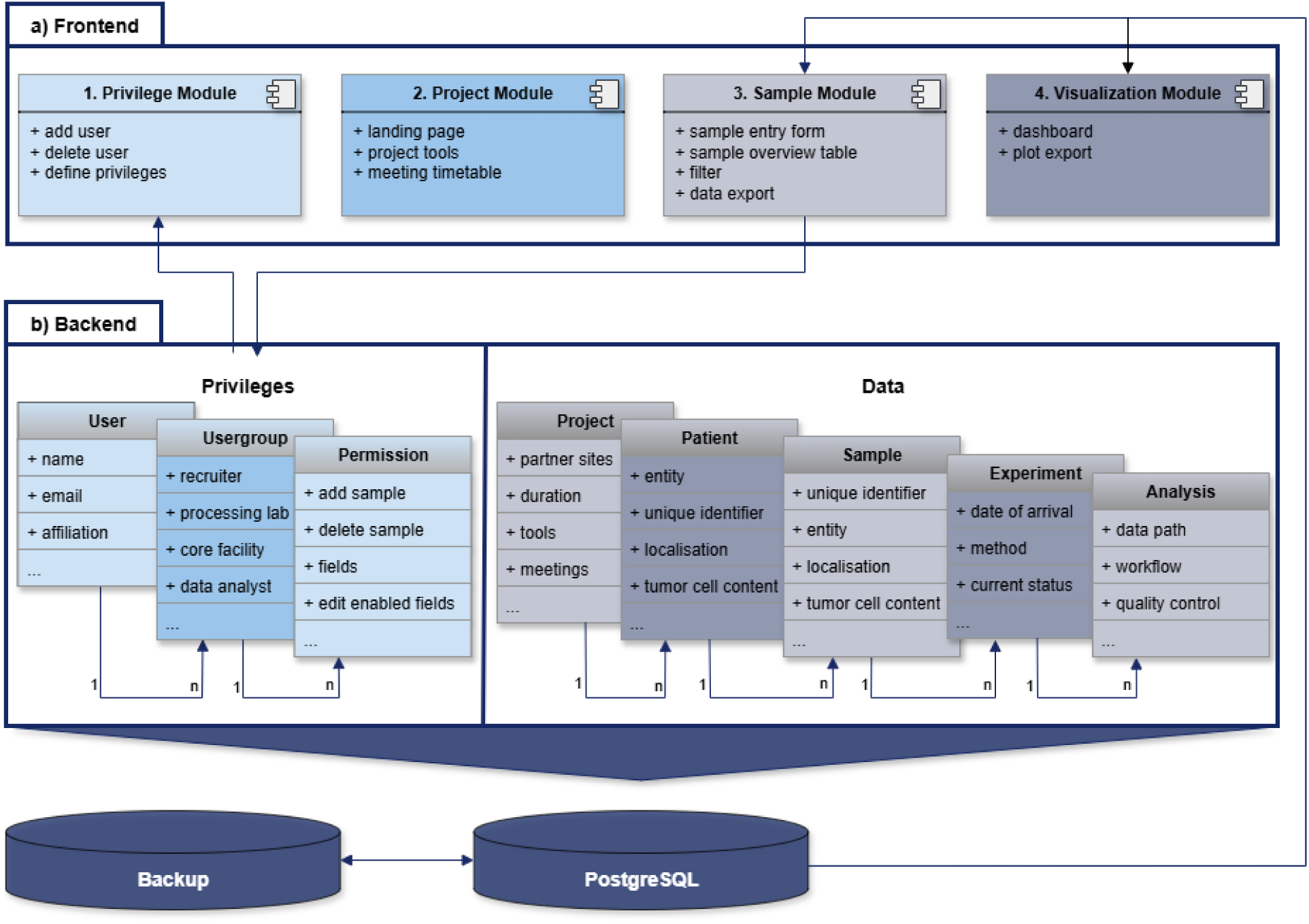
LiMeTrack application scheme. Overview of the LiMeTrack frontend modules (a) and the backend structure with underlying database (b).

Accordingly, the LiMeTrack backend keeps track of *Privileges*, *Data* and their details (Figure 2b). To enable fine-grained setting of user access privileges, *Permissions* for an individual *User* are tracked via *Usergroups* and correspond to either field-level permissions defining the database fields editable by that usergroup, or general permissions including the ability to add and delete samples. Similarly, a *Project* contains multiple physical patient-derived *Samples* identified by a unique identifier with customizable clinical annotations of entity, site, stage, grade, purity (in case of tumour samples) etc. For each *Sample* multiple *Experiments* are tracked, which correspond to the experimental analysis carried out, e.g. whole-genome sequencing or spatial transcriptomics. Each *Experiment* again might link to multiple instances of *Analysis*, which corresponds to one downstream bioinformatics analysis carried out on that specific sample (e.g. calling of different types of germline or somatic variants).

In the following we briefly describe a sample workflow from the SATURN3 project as processed in LiMeTrack across the main user group*s: Sample Recruiter*, *Sample Processor*, *Data Analysts* and *Project Manager*. The workflow starts with a biosample collection from a patient at a recruiting site. A unique patient pseudonym is given by an independent trust center. The *Sample Recruiter* adds the new biosample to the LiMeTrack *Sample Module* before sending it to the first *Sample Processor* being a sequencing facility involved in sample quality control and bulk sequencing. Project-relevant clinical information such as entity, age at diagnosis, localisation etc. are recorded at this stage as mandatory data. By using the *Sample Overview* or the *Dashboard* in LiMeTrack *Sample Processors* are aware of soon arriving samples and able to plan personnel and resources accordingly. Upon receiving the sample the a *Sample Processor* updates the LiMeTrack entry in the *Sample Module* by documenting arrival time and all following processing steps continuously. This documentation keeps all project partners updated in real time. For tissue samples a section is sent for pathological assessment and the pathologist documents tumor cell content and tissue quality in the *Sample Module*. A free text field allows for additional comments. Based on tissue quality and tumor cell content the decision for downstream analyses is taken. Each of the *Sample Processors* add metadata specific for their type of experiment which supports *Data Analysts* to conduct their analyses. Quality control (QC) reports can be linked in LiMeTrack accessible for all project partners who have granted read privileges and can download sample sheets specifically filtered for their needs. Finally, the *Dashboard* provides a succinct project overview and allows for the downloading of summary plots to be used in project-specific presentations and meetings. User and permission management is conducted by the *Project Managers* in the Admin section in the GUI.

Video 1 provides an overview of the platform and basic use cases.

### LiMeTrack allows for fine-grained permission management

As scientific biosample management in biomedical context depends on a high level of data security. Accordingly, all modules of LiMeTrack are password-protected and secured by state of the art security features of Django. A project manager who does not require programming knowledge can handle privilege management within a Django-based administrator interface. For SATURN3, 10 user groups exist with different read, write and editing privileges to defined data chunks stored within the platform. Write and editing access is only granted to data fields within the user group’s area of responsibility. This prevents all other fields from being accidentally filled or overwritten. For SATURN3 we allow only *Sample Recruiter* to add new biosamples to LiMeTrack and editing is enabled for selected *Project Managers* only. If necessary, individual permissions can be assigned on a per-user-basis in addition to the group-specific privileges. New user accounts can be easily created in single entry or batch format and by using the web application form or CSV file uploads.

### LiMeTrack efficiently tracks project and sample data

After login the LiMeTrack user is redirected to the project landing page which builds the core of the project module. Here all project related information and resources are linked and explained. A tabular overview lists regular project meetings with contact person and location or video call link. A change log keeps each user updated on newly implemented features and bug fixes. Based on the project module new colleagues can quickly familiarize themselves with the project during onboarding (Figure 3).

**Figure 3:**
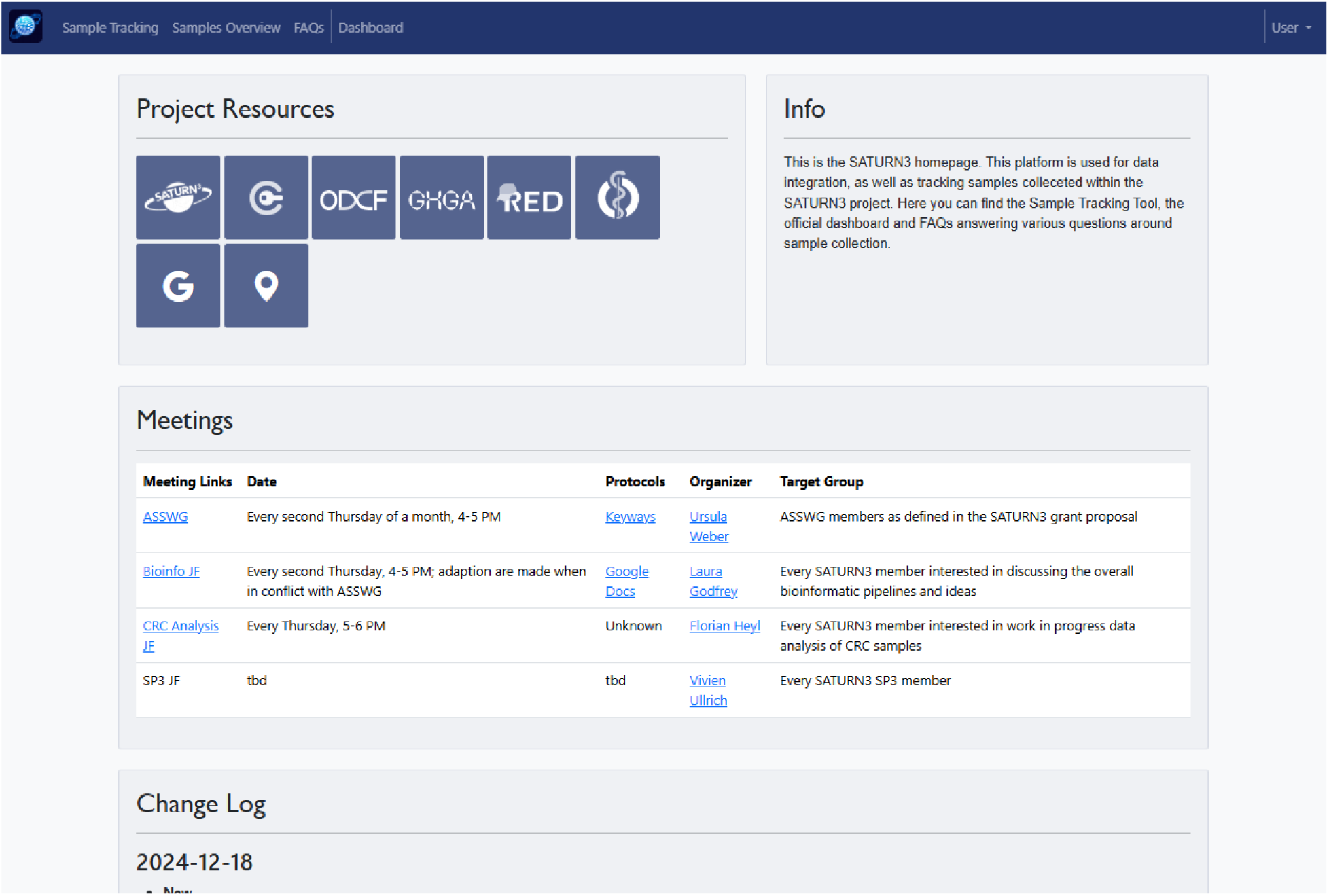
Exemplary LiMeTrack Project Module Page for the SATURN3 consortium. The project module serves as the landing page after login and displays an overview of all project related resources, important regular meetings as well as a change log of newly implemented features.

By clicking the *Sample Tracking* tab, the user enters the sample data entry mode, where data can be entered either as a single entry or via batch upload using CSV or Excel files. Specific template files are provided for download in the GUI. Explicitly defined entry restrictions (e.g., by dropdown menus or specified entry formats) ensure a high level of data consistency and minimize user errors, two common problems for multicenter projects where data is entered manually by different people. Before any data is uploaded to the database, a summary of all data entries is displayed and has to be confirmed by the user. In addition, internal checks validate the user entries, ensure data harmonization to a common format and allow the detection of inconsistencies. In case of any inconsistencies, clear and understandable error messages are prompted to indicate potential conflicts or missing data. The flexibility of the underlying programming infrastructure enables the fast and easy addition, removal, or adaptation of data fields to ensure flexibility with minimal programming knowledge. A categorization of the fields into mandatory and optional fields ensures that only complete data records can be entered. As an option, sequencing data quality control (QC) reports can be linked in a QC column to allow the connection of sample meta information with sequencing data. The current version of LiMeTrack actively used in the SATURN3 consortium consists of 61 data fields assigned to 10 different user groups. Table 1 summarizes the entry formats of the different fields. Free-text fields are kept to a minimum and do not involve any mandatory files but allow for free-text entry of notes and additional information that cannot be assigned to a distinct metadata field in specified structure.

**Table 1:** Summary of all data fields according to the respective category.

|  | **Dropdown** | **Restricted** | **Free-text** |
| --- | --- | --- | --- |
| **Number** | 18 | 38 | 5 |
| **Percent of all fields** | 29.5 | 62.3 | 8.2 |

### LiMeTrack provides real-time tabular and graphical data summaries and export options

The *Sample Overview* of LiMeTrack serves as the central data source available to every user and displays all data entries in a tabular form. To identify relevant subsets of samples, columns and rows can be filtered and a sample and patient counter in the header summarizes the number of total patients/samples in comparison to the applied filters. Furthermore, users could browse the sample table interactively based on distinct parameters by the provided search function of LiMeTrack. For support of various downstream applications, the whole sample table or the currently filtered subset can be downloaded in CSV or Excel format. In addition, LiMeTrack has implemented filters for bioinformatic workflows. These filters can be used to curate consistent sample sheets to execute computational analysis pipelines or to train models such as for bulk and single cell sequencing data. Each filter is designed to provide every covariate and data path variable needed for a specific workflow or model e.g. for variant calling. This reduces human errors and boosts the reproducibility and harmonization of the data processing and downstream analysis tasks.

Information in the database is used to create an interactive dashboard to ensure live and accurate project monitoring regarding processing status and project output (Figure 4). Interactive plots displayed in the dashboard can be exported in PNG format for presentations and project meetings. The dashboard can be easily extended to incorporate more figures based on the metadata stored within LiMeTrack.

**Figure 4:**
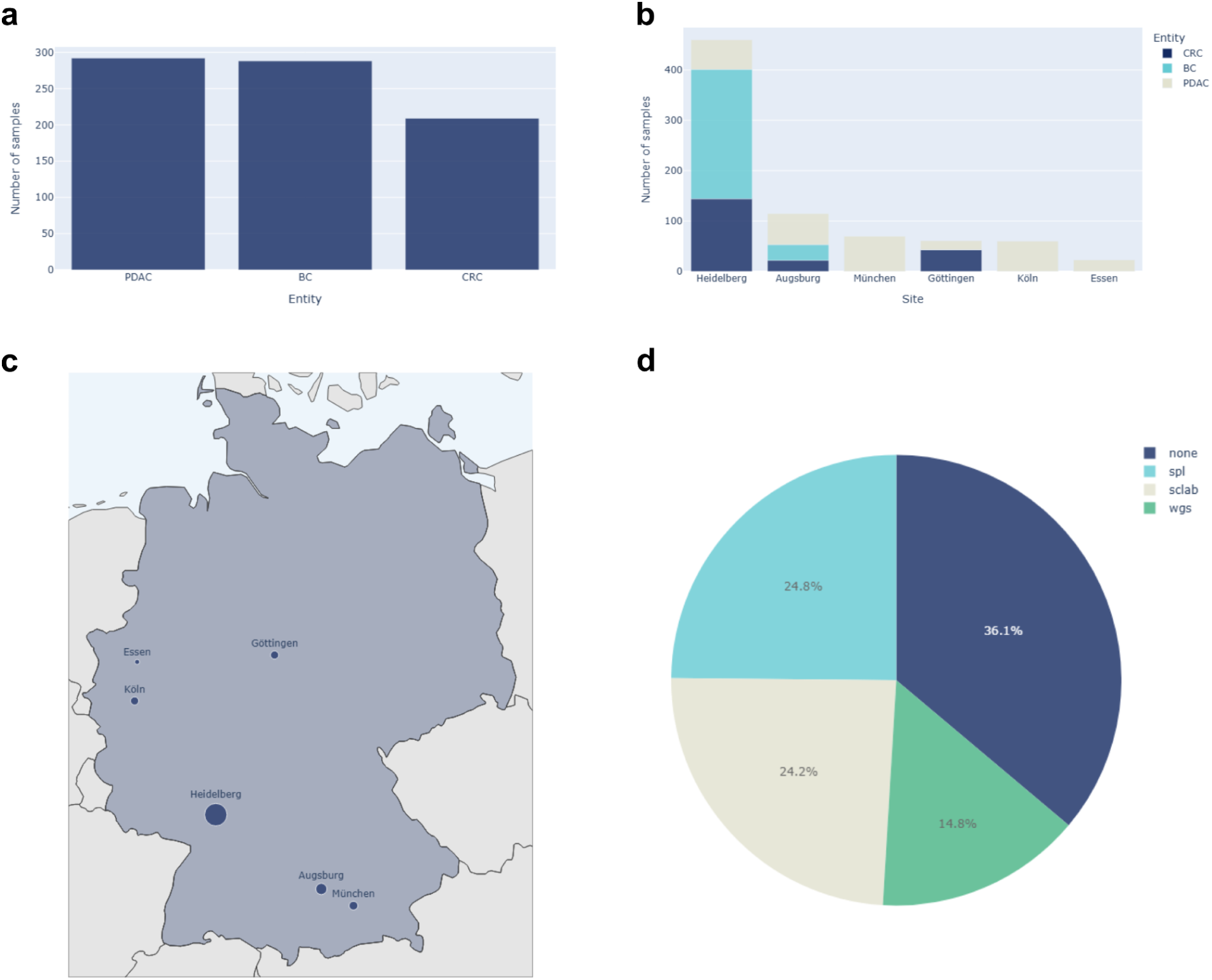
Exemplary LiMeTrack Dashboard. The dashboard plots display data of the underlying database based on study needs. a) Number of samples per entity are plotted. b) Bar graph representing the number of samples per entity for each recruiting site. c) Dot plot visualization of the sample number for each recruiting site on a map of Germany. The size of each dot corresponds to the number of samples documented in LiMeTrack. d) Pie chart representing the processing status of each sample. Abbreviations: BC: breast cancer, CRC: colorectal cancer, PDAC: pancreatic ductal adenocarcinoma, spl: sample processing lab, scLab: single cell processing lab, wgs: whole genome sequencing.

### LiMeTrack shows excellent usability in a real-world user study

To evaluate the practical usability of LiMeTrack in a real-world, multi-user setting, we conducted a user study with 23 participants (12 female, 11 male) based on the established System Usability Scale (SUS) developed by John Brooke [11]. The SUS score is a standardized, unitless measurement ranging from 0 to 100, which was later interpreted by Bangor et al. [12] to range from “worst imaginable” (0) to “best imaginable” (100), with a SUS score of 85.5 (sd=10.4) to mean “excellent”. Our results showed a mean SUS score of 81 (range 60-97.5) for LiMeTrack (Figure 5), indicating a high level of user satisfaction.

**Figure 5:**
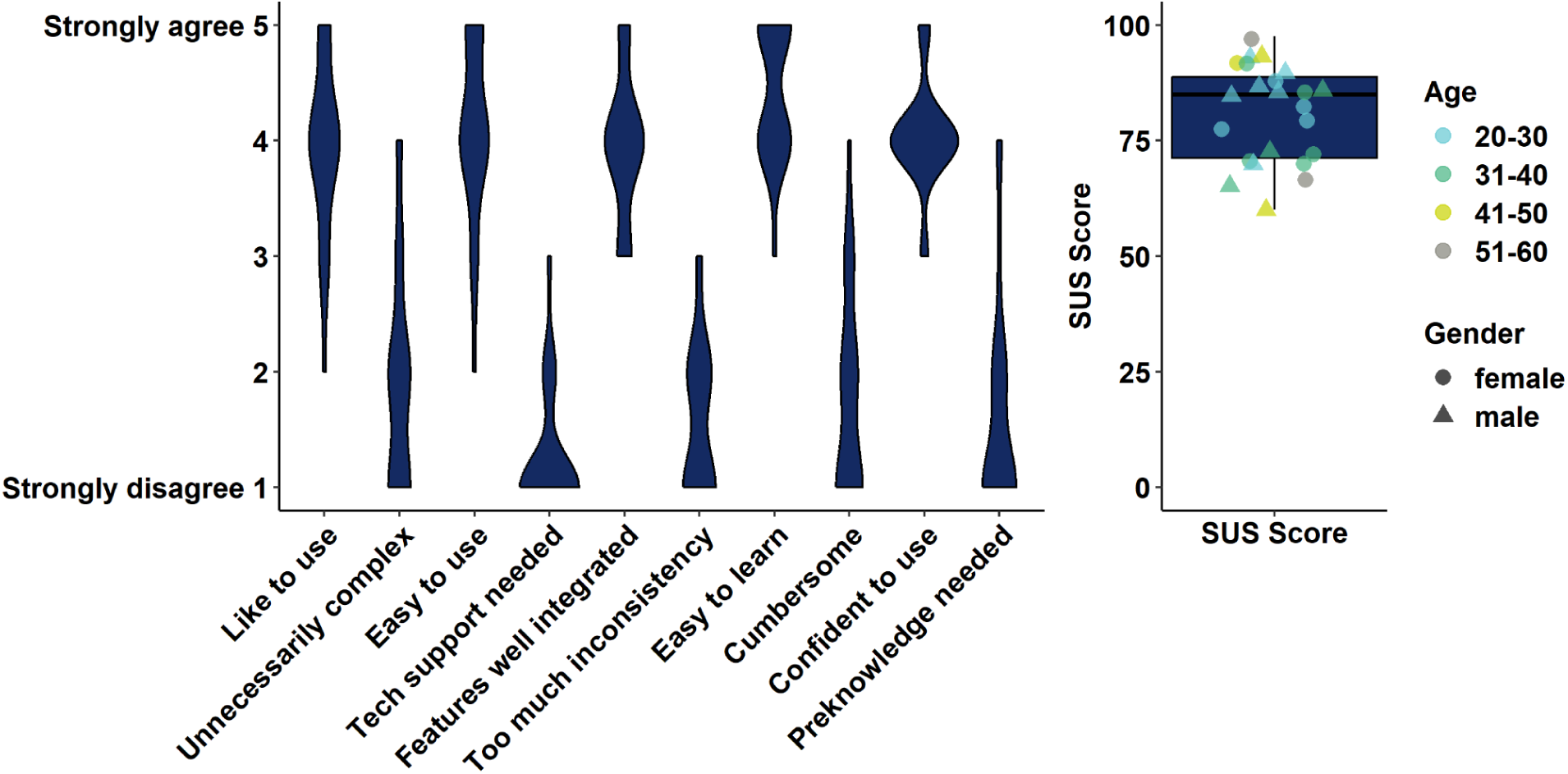
System Usability Scale. The user study was conducted with n=23 participants based on the questionnaire of John Brooke [11].

A SUS score of 70 and above traditionally meant passing a system usability testing and represented the mean SUS score in the evaluation of 959 SUS questionnaires by Bangor et al. [12]. Notably, 87% of participants rated LiMeTrack with a SUS score of 70 or above. None of the participants rated LiMeTrack unacceptable (SUS score below 50) further indicating LiMeTrack’s usability and appropriateness for biosample management.

The LiMeTrack GUI design focuses on efficient use especially for frequently performed tasks. Table 2 summarizes the number of mouse clicks needed for commonly performed user tasks. In particular, tasks not involving data entry can be mainly achieved with just one or two mouse clicks.

**Table 2:** Representative user tasks and mouse clicks needed to achieve. Mouse clicks are counted after login.

| User group | User task | Mouse clicks to achieve |
| --- | --- | --- |
| Recruiter | Enter new sample via entry form | 3 clicks<br>(fill of data entry fields excluded) |
| Recruiter | Enter new samples via batch upload | 4 clicks |
| Pathology | Add tumor cell content to existing sample | 5 clicks |
| Pathology | Add tumor cell content to existing sample via batch upload | 4 clicks |
| All | See total numbers of patients and samples | 1 click |
| All | Filter for patients of a specific entity | 2 clicks |
| All | Filter for patients of a specific entity and export filtered data table | 4 clicks |
| All | Export dashboard plot | 2 clicks |

Aiming for a platform that can be deployed and adapted with minimal programming knowledge we have emphasized well-structured and maintainable code. To quantitatively assess our codebase, we have measured various code metrics, as implemented in the Python module radon, v.6.0.1. In summary, LiMeTrack comprises 4873 lines of code (LOC) with 1834 logical LOC (LLOC). The average cyclomatic complexity of 169 code blocks was low (2.57) whereas the maintainability index score of all blocks was high (rank A).

## Discussion

In light of the increasing complexity of research projects involving patient biosamples, LiMeTrack provides a powerful all-in-one solution for sample tracking, data management, progress monitoring, and for serving analysis pipelines with relevant metadata. Its flexible and dynamic use allows for a direct and email-free communication between recruiters, wet labs, and analysts regarding the project status as well as sample and data quality. In this way, a clear project overview is ensured at all times and experiments and analyses can be planned in advance. A high level of user-friendliness was demonstrated in our user study. The flexibility of designing project-specific data entry fields, drop-down menus and user groups allows for a customization to individual project requirements.

Batch upload, validation, and error minimization reduce the workload of all project partners and lead to reliable research results. The dashboard overview ensures the efficient tracking of project progress and recruitment status therefore assisting project and budget management. In addition, structure and export options of the underlying database enabled the generation of respective metadata sheets to be used for data archives, e.g., Human Cell Atlas (HCA), German Human Genome Phenome Archive (GHGA), and European Genome Archive (EGA). In this way, an optimal re-use of the generated data is achieved, compliant with the FAIR principles [4]. LiMeTrack is designed to be used independently of the actual sequencing data storage, enabling the broadest possible application. By linking data storage paths and documents, LiMeTrack works with a wide variety of data management concepts and storage solutions. Our platform supports the generation of specifically tailored sample sheets of harmonized and structured metadata for various data analysis workflows. By combining sample tracking with automatic sample sheet creation, hands-on time for manual data curation is reduced. Furthermore, mandatory entry fields ensure availability of all relevant metadata. This streamlined process not only improves productivity but also minimizes the risk of human error. In the current version of LiMeTrack, workflows for analyses of bulk (e.g., nf-core/sarek) and single-cell sequencing (e.g., nf-core/scrnaseq) as well as spatial transcriptomics (e.g., nf-core/spatialxe) are implemented and are successfully applied in the SATURN3 consortium. Thus, LiMeTrack already supports a broad range of data modalities while the flexible platform design allows for a fast extension to project specific data types and pipelines.

Overall, LiMeTrack addresses a plethora of potential failure points encountered when using spreadsheets for sample tracking in multimodal and interdisciplinary research projects. In the SATURN3 project we have reached a state of consistency far beyond shared spreadsheets without losing the ease of use or comfortability valued by most users. LiMeTrack has equipped us with the flexibility to fulfill the evolving needs of a dynamic research project. This enabled the seamless addition of new data entry fields for e.g. new modalities or the implementation of filters for new types of analysis workflows.

There remain potential limitations. Despite the lightweight computational framework for modification, programming skills especially to customize the dashboard and underlying database are still required. Also the deployment has to be realized on a project-specific basis. We are working on further simplifying the data models to be modularly adaptable for a wide range of projects, e.g., involving patient-derived organoid models. In the future, the extension of the current tool stack is planned. We will mainly focus on APIs for commonly used interoperability workflows to connect sample metadata with sequencing results and imaging data, such as LaminDB [13] or SpatialData [14]. Furthermore, we will further evaluate our entry restrictions in practical use and further optimize internal data validation and error minimization strategies. These ongoing developments will further enhance LiMeTrack’s functionality and strengthen its role as a valuable infrastructure component for advancing large-scale biomedical research projects.

## Methods

### LiMeTrack implementation

LiMeTrack is a web platform built using the modern Python-based and widely used Django web framework that allows for a secure, rapid development and high flexibility (Django Software Foundation [15]). All data entered via the front-end is persisted in a tailored PostgreSQL-database.

Due to the microservice nature, LiMeTrack provides the scalability and flexibility needed when entering or managing large amounts of data as well as the extensibility to implement project specific features. Using Django for traffic routing and the user interface, together with its administrative capabilities allows for fine-grained privilege and user management. Currently, LiMeTrack distinguishes between read-only permissions, data entry permissions as well as editing and deletion permissions. All can be set in a modular fashion for user groups and/or individual users.

The LiMeTrack infrastructure enables various access methods, for example direct database access for data engineers, and is compatible with other applications by direct database access or extending it with the Django REST framework.

The containerized application as well as the database are hosted on a dedicated virtual machine (VM) behind an application firewall on premises of the University Clinic Freiburg and deployed using the Docker tech stack [16]. The risk of data-loss is minimized through a well planned backup strategy involving doing hourly incremental and nightly full backups. Increased debugging capabilities and data-protection conform traceability are realized through extensive logging. Front-end access as well as all CRUD (Create, Read, Update, Delete) operations to the database are registered and also backed up.

### User study

To assess usability, 23 volunteers (12 female, 11 male) were recruited through snowball sampling. Following a 5-minute introduction and live demo of LiMeTrack’s main features, the participants were asked to assume the role of a recruiter. Two samples per person were given to be entered via the submission form. In addition, an Excel spreadsheet containing two additional samples was provided but contained a non-valid value for sex and an incorrectly formatted patient pseudonym. Study participants were asked to identify and correct the errors. Finally, three questions should be answered using the filter functions and the dashboard. Afterwards, feedback was collected using the System Usability Scale (SUS) questionnaire as proposed by John Brooke [11]. The System Usability Score was calculated accordingly.

## Data Availability

LiMeTrack source code and a standalone Docker image are available on Bitbucket as a GIT repository (https://bitbucket.org/schwarzlab/limetrack).

## Data and code availability

LiMeTrack source code and a standalone Docker image are available on Bitbucket as a GIT repository (https://bitbucket.org/schwarzlab/limetrack). Setting up and monitoring the service is made straightforward with a docker-compose file as described in the documentation.

## Declarations

OpenAI (2023). ChatGPT (GPT-4, GPT-4o) [Large language model] was used for text refinement and copy editing of abstract, introduction and discussion [17].

## Author Contributions

Conceptualization, FH, JG, FV, LH, RFS, LG

Methodology, FH, JG, FV, LH, LG

Software, JG, FV, LH

Writing – Original Draft, FH, RFS, LG

Writing – Review & Editing, all authors

Visualization, LG

Supervision, MB, RFS, LG

Funding Acquisition, OS, JK, JTS, MB, RFS

### Acknowledgements

We thank Daniel Schütte (ICCB Cologne) for advice on system usability metrics and critical review of the manuscript and Alexander Nicolay (ICCB Cologne) for supporting video recording and processing.

